# The involvement of male partners in care: socio determinants of health among women in Prevention of Mother to Child Transmission in Malawi

**DOI:** 10.1101/2020.04.18.20070714

**Authors:** Isotta Triulzi, Olivia Keiser, Claire Somerville, Sangwani Salimu, Fausto Ciccacci, Ilaria Palla, Jean Baptiste Sagno, Jane Gondwe, Cristina Marazzi, Stefano Orlando, Leonardo Palombi, Giuseppe Turchetti

**Author notes:** **Corresponding author’s name** Isotta Triulzi, Piazza Martiri Libertà, 567126 Pisa (Italy).

## Abstract

**Introduction:** Male partners are rarely present during PMTCT (Prevention-Mother-To-Child-Transmission) services in Sub-Saharan Africa (SSA). The involvement of men is increasingly recognised as an important element of women’s access to care. The study aims to identify the socio-demographic characteristics, HIV-Knowledge, Attitude and Practice (KAP) among women accompanied and not accompanied by the male partners to the facilities.

**Method:** We included pregnant women enrolled in PMTCT programme between August 2018 and November 2019 in the Southern Region of Malawi. Eligible women were aged 18 years or older, lived with a male partner, enrolled for the first time in four DREAM facilities. We provided a KAP survey to women and their partners attending the facilities. Our primary outcome was to assess and analyse the proportion of women who were accompanied by their partner at least once. We applied descriptive statistics, and logistic regressions to study the association between being accompanied and explanatory variables.

**Results:** We enrolled 128 HIV-positive women : 82 (64.1%) were accompanied by their male partners and 46 (35.9%) were alone. Women with high level of knowledge on HIV/AIDS are more likely to be accompanied by the male partners (53.7% vs 34.8%,p=0.040). Level of attitude and practice were not different between women accompanied or not. Patients owning a mean of transport were more likely to go alone to facility (OR 4.16, 95% CI 1.02-16.94). Women who travelled more than 90 minutes to get to the facilities (OR 0.10, 95% CI 0.02-0.49) with high HIV-knowledge (OR 0.38, 95% CI 0.16-0.91) are more likely to be accompanied.

**Conclusion:** Our study showed a good male partner involvement compared to other studies performed in SSA. To our knowledge this is the first study outlining the relationship between male partner involvement in care with socio determinant of health. This is crucial to design and implement effective interventions.

## Introduction

In 2018, 37.9 million people across the globe were living with HIV/AIDS. Of these, 36.2 million were adults and 1.7 million were children (<15 years old). The WHO African Region was most severely affected with 25.7 million people living with HIV. Worldwide, there were 1.3 million pregnant women living with HIV in 2018, of which 82% received Antiretroviral Therapy (ART) for Prevention Mother-To-Child Transmission (PMTCT) of HIV (World Health Organization 2019).

Malawi is one of the countries with higher HIV prevalence in the adult population (15–49 years) amounting to 9.2% in 2018 (United Nation AIDS n.d.). It is estimated that one million Malawians, adults and children, are infected with HIV. Women infected represent 59.8% of the total adult population.

The literature shows that in Sub-Saharan Africa (SSA), male partners are scarcely present during PMTCT services (World Health Organization 2012). In many low-income countries, the involvement of men in maternal services is increasingly recognised as an important element of women’s access to needed care (Tokhi et al. 2018; World Health Organization 2015). Male partner involvement is not a well-defined concept and there is currently no single widely used indicator to measure it. Several definitions have been used in previous studies (Byamugisha et al. 2011; Farquhar et al. 2004; Kalembo et al. 2013), but the most comprehensive one was proposed by Muwanguzi et al (Muwanguzi et al. 2019). In this study male partner involvement in PMTCT was not limited to accompany the woman to the clinic and perform Couple HIV Testing and Counselling (CHTC), but also to support the woman during the treatment from an economic and psychosocial point of view. Some studies demonstrated that the male partner support is relevant to the adherence to therapy of women in PMTCT. The study performed by Katirayi et al. (Katirayi et al. 2017) in Zimbabwe and Malawi shows that one of the main barriers to women initiating and adhering to ART were their male partners. Zacharius et al. (Zacharius et al. 2019) showed that women with male support in PMTCT were 3.5 times more likely to have good adherence than those without support. Male involvement has been recognized as a priority in PMTCT but currently remains a challenge in most low- and middle-income countries. The literature explains the barriers (Cuco et al. 2015; Morfaw et al. 2013), few programs have been already implemented, but still much improvement is needed to enhance the partner male involvement, thus it is important to identify better strategies to involve male partners in PMTCT and in healthcare services (Word Health Organization 2014).

The paper represents the first stage of a study aiming to identify interventions to improve male involvement along the cascade of HIV services in Malawi. This paper aims to: 1) assess the involvement of male partners in PMTCT; 2) describe the socio-demographic characteristics of women enrolled in PMTCT, accompanied by their male partner or not; 3) determine the level of knowledge, attitude and practice toward HIV/AIDS associated with male accompanied; 4) study the association between being accompanied by a male partner and socio-demographic characteristics and knowledge attitude and practice on HIV/AIDS.

## Methods

### Study population and setting

We included pregnant women enrolled in Malawi’s national PMTCT programme between 10 August 2018 and 30 November 2019 at four health facilities in the Southern region of Malawi. These health facilities are part of the DREAM (Disease Relied through Excellent and Advanced Means) programme sub recipient of Global Fund, a public health program run by the Community of Sant’Egidio in 11 African Countries focusing on HIV/AIDS, TB and NCDs (Ciccacci et al. 2019; Marazzi et al. 2014; Orlando et al. 2018). The facilities are located in Blantyre (urban), Machinjiri (peri-urban), Chileka (peri-urban) and Balaka (rural).

The pregnant women were eligible if they were aged 18 years or older, lived with a male partner, enrolled for the first time in a DREAM facility and they were willing to sign a written informed consent.

The study is a part of a larger research program that aims to investigate the impact of male participation on women’s ART adherence in PMTCT in Malawi.

We provided a Knowledge, Attitude and Practices (KAP) survey (World Health Organization 2008) to women and their male partners attending the health facilities. KAP surveys assess knowledge gaps, cultural beliefs, and behavior of people with HIV/AIDS. They identify misconceptions or misunderstandings that may represent barriers to accessing health care, the uptake of interventions and the improving of adherence to ART. We designed the survey questionnaire following the guideline for conduction a KAP survey (World Health Organization 2008, 2014) The final questionnaire consisted of 27 questions or statements (see Appendix): 15 on knowledge of HIV transmission (1-8), prevention and anti-retroviral therapy (9-15), 7 on attitudes towards HIV/AIDS, and 5 on sexual, alcohol or drug use behaviour. The answer to each question had three categories (yes, no, do not know).

We discussed the readability and clarity of the questionnaire with two healthcare workers and a medical doctor to better adapt the questions to the Malawian context. We then pilot-tested it in a small group of randomly selected women (n=22). The KAP survey was administered to a woman and her male partner the first time she or he came to the clinic. If the male partner did not attend any clinical visit during the study period, the questionnaire was completed by the woman only, within one month and two weeks after the first visit. It is recommended that women have at least three PMTC visits after the first visit.

### Outcomes

The primary outcome was the proportion of women in PMTCT who were accompanied by their male partner at health facilities at least once during a follow-up period of six weeks. Among the women accompanied we collected secondary male partner specific outcomes, as the proportion of women who i) disclosed their status to their male partner, ii) reported to their male partners that they had a clinical appointment, iii) asked the male partner’s permission before going to the facilities, iv) who delivered the invitation slip and v) the proportion of women who received money by the male partner for the transport to the clinic. Our primary outcome is the baseline requirement for considering the male partner involved in PMTCT. The secondary outcomes were reported by the women not accompanied by the partner.

In addition to the KAP survey we also used data from the electronic medical record system on the women enrolled in the study. Data included age, educational level, type of job, number of people in the family, socio-economic condition (e.g. availability of electricity), means of transport and distance to the facility. A woman was considered lost to follow-up (LTFU) when she missed to collect the doses of antiretroviral drugs by more than two months.

## Statistical analyses

The characteristics of the women were described using Fisher test or chi-square tests for categorical variables, and Wilcoxon rank sum tests for continuous variables.

We determined the percentage of correct answers for KAP question, similarly to previous studies (Nubed and Akoachere 2016; Shokoohi, Karamouzian, and Mirzazadeh 2016; Thanavanh, Kasuya, and Sakamoto 2013). The answers to the KAP questions were scored: a score of 2 was attributed for a correct answer, a score of 1 for an uncertain answer and a score of 0 for a wrong answer. The scores were summed to develop an overall score for each woman. We compared the score using Mann-Whitney tests. We then determined the median score for each of the three sections in order to determine if there was any difference in knowledge, attitude and practices among women accompanied and not accompanied by their male partner. We compared the knowledge, attitude and practices using Mann-Whitney tests with 95% confidence intervals (CI).

We conducted univariable and multivariable logistic regressions to study the association between being accompanied by a male partner (yes, no) and explanatory variables. Explanatory variables included age (18-25.5, >25.5 years), education (no education/primary school, secondary school/pre-University), employment (houseworker, employee, unemployed, temporary job), availability of transport including cars, motor cycles and bikes (yes, no), time to the facility (0-60 minutes, 61-90 minutes, >90 minutes), economic condition (availability of electricity in household, yes, no), and knowledge (low, high), attitude (positive, negative), practice (safe, risky) toward HIV. The level of knowledge, attitude and practice was categorized using the respective median as cut-off, as suggested by Thanavanh et al (2013).

We imputed missing values of explanatory variables by using multiple imputation with chained equations (MICE). We added the following variables to improve the imputation (Sterne et al. 2009): mother alive (yes/no), owns a phone (yes/no), gestational age (1-3, 4-6, >7), piped water available in the dwelling (yes/no), type of transport (bus, others). Furthermore, we considered also the outcome in the imputation. Given the low sample size we run the model with six explanatory variables: age, education, mean of transport, time to the facility, employment status and level of knowledge on HIV/AIDS. Age and education were included a priori as relevant demographic factors, the other variables gave significant results in the univariable logistic regressions. We ran the model on 20 imputed datasets for each analysis and combined the estimates with Rubin’s rule (Rubin 2004).

All analyses were performed using STATA 13.

### Ethical consideration

The patients were informed about the objectives of the project and written informed consent was obtained from all participants. The study protocol has been approved by the National Health Sciences Research Committee (Minister of Health) in Malawi [approval number 2021].

## Results

### Characteristics of study participant and KAP survey

We screened 142 HIV-positive pregnant women. A total of 128 women met the eligibility criteria: 82 (64.1%) were accompanied by their male partners and 46 (35.9%) were alone. Two women came with their partners before their transfer to another facility. No women were lost to follow-up during the study. Table 1 shows the socio-demographic characteristics of the participants, and they knowledge, attitude and practice toward HIV in the male partner-accompanied and in the male partner-non-accompanied group. The median age of the women was 27.8 years (IQR 22.8-32.3) with no significant difference in the two groups. Half of the women had no education or attended the primary school (46.8%). Significant difference in terms of employment, owning a mean of transport, travel time to facilities were found among the two groups. Accompanied women were more likely to be houseworkers than non-accompanied women (75.6% vs 56.5%, p=0.045) and had a mean of transport in a lower number of cases (6% vs 23.9%, p=0.003). Among the 28 women who took more than 90 minutes to get to a facility, the majority (89.3% vs 10.7%, p=0.000) were accompanied by the male partner. In term of economic condition and size of family no significant differences in the two group were found.

**Table 1.** Socio-demographic information of HIV-positive pregnant women in PMTCT in Malawi (*n*= 128)

|  | All (n=128) | Female accompanied by the male partner (n=82) | Female not accompanied by the male partner (n=46) | P-value* |
| --- | --- | --- | --- | --- |
| **Socio-demographics variables** |  |  |  |  |
| **Age** Median (IQR) | 27.8 (22.8-32.3) | 27.4 (22.0-32.3) | 28.6 (24.6-32.3) | 0.356 |
| **Age** |  |  |  |  |
| 18-25 | 50 (39.1) | 33 (40.2) | 17 (37.0) | 0.902 |
| >25 | 78 (60.9) | 49 (59.8) | 29 (63.0) |  |
| **Educational level** |  |  |  | **0.823** |
| No education/ Primary | 60 (46.9) | 39 (47.6) | 21 (45.6) |  |
| Secondary/Pre-University | 65 (50.8) | 41 (50.0) | 24 (52.2) |  |
| Missing | 3 (2.3) | 2 (2.4) | 1 (2.2) |  |
| **Healthcare center location** |  |  |  | **0.302** |
| Machinjiri | 74 (57.8) | 36 (43.9) | 38 (82.6) |  |
| Chileka | 35 (27.3) | 31 (37.8) | 4 (8.6) |  |
| Balaka | 12 (9.4) | 10 (12.2) | 2 (4.4) |  |
| Blantyre | 7 (5.5) | 5 (6.1) | 2 (4.4) |  |
| **Employment Status** |  |  |  | **0.045** |
| Houseworker | 88 (68.8) | 62 (75.6) | 26 (56.5) |  |
| Unemployed | 3 (2.3) | 1 (1.2) | 2 (4.3) |  |
| Employee | 9 (7.0) | 5 (6.1) | 4 (8.7) |  |
| Temporary job | 22 (17.2) | 9 (11.0) | 13 (28.3) |  |
| Missing | 6 (4.7) | 5 (6.1) | 1 (2.2) |  |
| **Partner's Employment** |  |  |  | **0.359** |
| Employee | 45 (35.2) | 25 (30.5) | 20 (43.5) |  |
| Unemployed | 70 (54.7) | 48 (58.5) | 22 (47.8) |  |
| Missing | 13 (10.1) | 9 (11.0) | 4 (8.7) |  |
| **Size of your family** |  |  |  | **0.280** |
| 0-3 | 31 (24.2) | 23 (28.0) | 8 (17.4) |  |
| >3 | 96 (75.0) | 59 (72.0) | 37 (80.4) |  |
| Missing | 1 (0.8) | 0 | 1 (2.2) |  |
| **Travel time to facility (minutes)** |  |  |  | **0.000** |
| 0-60 minutes | 61 (47.6) | 41 (50.6) | 20 (44.4) |  |
| 61 - 90 minutes | 37 (28.9) | 15 (18.5) | 22 (48.9) |  |
| >90 minutes | 28 (21.9) | 25 (30.9) | 3 (6.7) |  |
| Missing | 2 (1.6) | 1 (1.2) | 1 (2.2) |  |
| **Having a mean of transport** |  |  |  | **0.003** |
| Yes | 16 (12.5) | 5 (6.0) | 11 (23.9) |  |
| No | 109 (85.2) | 75 (91.5) | 34 (73.9) |  |
| Missing | 3 (2.3) | 2 (2.5) | 1 (2.2) |  |
| **Electricity available in the dwelling** |  |  |  | 0.167 |
| Yes | 77 (60.2) | 29 (35.4) | 22 (47.8) |  |
| No | 51 (39.8) | 53 (64.6) | 24 (52.2) |  |
| **Knowledge, attitude and practice toward HIV** |  |  |  |  |
| **Knowledge** |  |  |  | **0.040** |
| Low | 68 (53.1) | 38 (46.3) | 30 (65.2) |  |
| High | 60 (46.9) | 44 (53.7) | 16 (34.8) |  |
| **Attitude** |  |  |  | 0.214 |
| Negative | 52 (40.6) | 30 (36.6) | 22 (47.8) |  |
| Positive | 76 (59.4) | 52 (63.4) | 24 (52.2) |  |
| **Practice** |  |  |  | 0.836 |
| Risky | 68 (53.1) | 43 (52.4) | 25 (54.3) |  |
| Safe | 60 (46.9) | 39 (47.6) | 21 (45.7) |  |
| **Total** | 128 (100) | 82 (100) | 46 (100) |  |

\* Chi-square or Wilcoxon rank sum test

Responses to questions on the KAP survey are reported in the Supplementary (Tables 1,2,3). The analysis of the KAP survey showed that women with high level of knowledge on HIV/AIDS were more likely accompanied by the male partners (53.7% vs 34.8%, p=0.040), instead level of attitude and practice toward HIV were not different in the two groups (Supplementary, Table 4). No association was found between the level of attitude and practice toward HIV among accompanied and not-accompanied women.

**Table 2.**
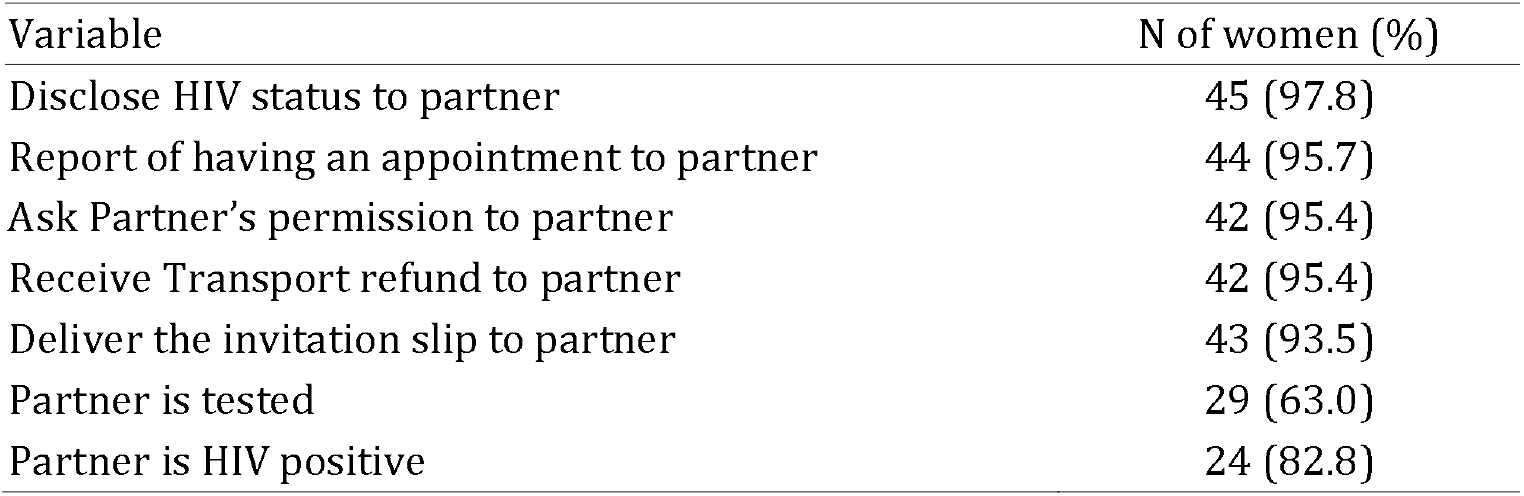
“Male partner specific” variables among women not accompanied (n=46)

**Table 3.** Univariable and multivariable analyses examining association between HIV positive pregnant women being accompanied or not by the male partners to the health facilities in Malawi (n=128)

|  | Crude OR<br>Estimate (95% CI) | Adjusted OR<br>Estimate (95% CI) |
| --- | --- | --- |
| Socio-demographic variables |  |  |
| Age (years) |  |  |
| 18-25 | 1 | 1 |
| >25 | 1.15 (0.55-2.42) | 0.83 (0.37-1.90) |
| Education |  |  |
| No education/primary | 1 | 1 |
| Secondary/Pre-University | 1.09 (0.52-2.26) | 0.83 (0.36-1.89) |
| Employment |  |  |
| Houseworker | 1 | 1 |
| Unemployed | 4.84 (0.42-55.75) | 0.53 (0.02-14.01) |
| Temporary job | 3.48 (1.33-9.13) | 2.51 (0.79- 7.9) |
| Employed | 1.94(0.48-7.79) | 3.72 (0.62-23.04) |
| Mean of transport |  |  |
| Yes | 1 | 1 |
| No | 4.12 (1.01-15.06) | 4.16 (1.02-16.94) |
| Travel time to facility (minutes) |  |  |
| 0-60 | 1 | 1 |
| 61- 90 | 2.91 (1.25-6.76) | 1.48 (0.62-3.53) |
| >90 | 0.24 (0.70- 0.90) | 0.10 (0.02-0.47) |
| Electricity available in the dwelling |  |  |
| No | 1 |  |
| Yes | 1.68 (0.80- 3.49) |  |
| Knowledge, attitude and practice toward HIV |  |  |
| Level of Knowledge |  |  |
| Low | 1 | 1 |
| High | 0.46 (0.22- 0.97) | 0.38 (0.16-0.91) |
| Level of Attitude |  |  |
| Negative | 1 |  |
| Positive | 0.63 (0.30- 1.31) |  |
| Level of Practice |  |  |
| Risky | 1 |  |
| Safe | 0.93 (0.45 - 1.91) |  |

OR = odds ratio; CI = confidence interval

Table 2 shows “male partner specific” variables among the women not accompanied by the partner. Twenty-nine (63%) of men tested for HIV and 24 (82.8%) were HIV positive. The median age of the partners was 35.5 years (IQR 32-38) and 39 (84.8%) were employed.

Several sociodemographic variables were associated with going alone to the health facility in the univariable analysis (Table 3). These included having a temporary job (odds ratio [OR], 3.48; 95% confidence interval [CI], 1.33-9.13), owing a mean of transport (OR 4.12; 95% CI, 1.01-15.06), living between 60 to 90 minutes (OR 2.91, 95% CI 1.25-6.76) from the health facilities. Also, women with high level of knowledge (OR 0.46, 95% CI 0.22-0.97) were more likely to go alone in the univariable analysis.

In the multivariable model, owning a mean of transport, living more than 90 minutes from the health facility and demonstrated a high level of knowledge were associated with the outcome. Patients owning a mean of transport were more likely to go alone to the facility (OR 4.16, 95% CI, 1.02-16.94). Women who travelled more than 90 minutes to get to the facility (OR 0.10, 95% CI 0.02-0.47) and with a high knowledge on HIV (OR 0.38, 95% CI 0.16-0.91) were more likely to be accompanied by the male partner.

## Discussion

In this study we investigated the male partner involvement in a sample of women included in a PMTCT program in Malawi. We evaluated the association between the involvement of the male partners in care and the socio-demographic characteristics, HIV-knowledge, attitudes and practices related to HIV/AIDS. To our knowledge this is the first study comparing the socio-economic characteristics and knowledge, attitudes and practices of women accompanied and not accompanied to the healthcare facility by the partner.

Several studies evaluate the male partner involvement. Our study shows that the male partner involvement is 64.1% in PMTCT. This is comparable to the result reported by Rosenberg et al. (2015) (Rosenberg et al. 2015) in a unblinded randomised controlled trial comparing the adoption of the invitation slip versus the use of invitation slip plus tracing (52% vs 74%) in Lilongwe, Malawi. Other studies performed in Malawi reported that the male attendance at antenatal clinic were: 13.7% in a retrospective study performed in 2004-2006 in Mwanza District (Kalembo et al. 2013), 10.7% in a observational study over three periods where the researchers introduced peer education and designed a male-friendly infrastructure in Bwaila Hospital in Lilongwe in 2009 (Mphonda et al. 2014), 19% in a randomized control trial where the use of an invitation slip was compared to non-intervention (Nyondo et al. 2015). In other countries, the trials suggested that the male attendance ranged from 16% in Uganda (Byamugisha et al. 2011), 35% in South Africa (Mohlala, Boily, and Gregson 2011), 36 % in Kenya (Osoti et al. 2014) to 53.5 % in Tanzania (Jefferys et al. 2015). Our result shows a good male partner involvement compared to other studies. One reason could be that in the last few years more attention and more interventions have targeted male involvement in the country.

Our study shows that the distance from the healthcare facility and the possession of a vehicle were associated with male accompaniment. Both of these results reveal the importance of the social determinants of health (Schulz and Northridge 2004). Owning, or having access to a vehicle does not only indicate easier access to the healthcare facility but it is strongly associated with higher income and social class, especially in Malawi that is one of the poorest countries in the world where “poverty and inequality remain stubbornly high” (Anon 2019). Distance to healthcare facility is still indicated by 56% of women as a key barrier to health access when they are sick (Anon 2016) with a median travel time of 1 hours (Varela et al. 2019). In the future it would be interesting to consider the geo-distribution (Maina et al. 2019) of the population and distance to facilities in our sample. Moreover, the extreme poverty of the country astonished us when we see that 51.5% of the population income fell below the poverty line in 2016 (Anon 2019). A recent published research showed that 56% of household would spend less than 1 US$ to reach the healthcare facility and that in the country are present only 14.3 cars and motor cycles per 1000 people (Ministry of Transport and Public Works 2019). In fact, lack of financial resources to go to a hospital has been reported by 39% of male and 59% of female head of households as one of the main barriers (Varela et al. 2019).

A relevant result is that a higher score in the level of HIV-knowledge was associated with a greater involvement of the male partner, but attitudes and practices were not associated with outcome. This last aspect reveals a limit of health education as a way of generate a behavioural change; as other scholars already reported, knowledge does not always turn into different attitudes or practices (Kok, van den Borne, and Mullen 1997). The low level of education of the study participants certainly plays a role (Van Der Heide et al. 2013; Pandit et al. 2009); overall 46.8% of the women interviewed did not receive any education or only attended primary school. The National Statistical Office (National Statistical Office (NSO) [Malawi] 2017) reported differences in access to education by sex: 81% of women and 73% of men have never attended school or have some primary education all over the country reflecting wider structural inequalities including gender. In terms of gender, male-headed households are richer that female-headed households (43% vs 57 % of the total households) (International Monetary Fund. 2017) that may indicate a lower access to education. Moreover, even if the level of education among men is slightly higher than women, this study confirms findings in many other studies that health education targeting only women is important, but it is not enough to increase the involvement of their male partners. Interventions that target directly the male partners and that adopt an intersectionality approach to include geographical accessibility of health centres, income, social class, social practices and gender are needed (Hankivsky 2012; Somerville 2020).

This study has some limitations. Firstly, the limited size of the sample prevented stratifications in statistical analysis and reduced the power of some analyses. Secondly, due to the non-randomisation some biases occurred, for example the women who participated may be more involved in the program or more informed. Thirdly, attitudes and practices were self-reported incurring social desirability bias (Launiala A 2009). For this reason, in future studies we intend to evaluate objective outcomes, such as viral load suppression (adherence to ART). Fourthly, there are no standards for calculating scores in a KAP analysis that allow us to compare our results with others in a similar context. However, the majority of KAP questions are also present in the Demographic and Health Surveys (DHS) performed in several Sub-Saharan African countries by USAIDS (United Stated Agency International Development USAID n.d.). A direct comparison was nevertheless not possible due to the different sampling method.

Our study suggests that the involvement of male partners could be improved by considering other socio-determinants of health beyond only health education. Interventions with integrated multi-component strategies (Triulzi et al. 2019) delivering healthcare services closer to the villages or at home, or the use incentives (economic or non-economic) may increase male participation (Besada et al. 2016; Office of Population and Reproductive Health Bureau for Global Health 2018). As shown by Salmen C.R. et al. (Salmen et al. 2015) promoting social network engagement through the development of “microclinic” intervention in villages (consists of a small network of 5-15 neighbours, relatives and friends) may be promising and could be designed to evaluate the involvement of male partners. In order to design effective intervention to improve male partner engagement we have to recognise wider Malawian gender orders and gender norms around masculinities (Chikovore et al. 2015). Such “restrictive gender norms” (Hay et al. 2019) and the wider societal inequality regimens (Acker 2006) are reproduced in the healthcare systems (Kanter and Stein 1979), Somerville, 2020 forthcoming) and may impact male partner engagement. As Gary Darmstadt and colleagues showed in the Lancet Series on Gender Equality, Norms, and Health, gender norms and inequities are determined and reinforced in families, communities, structures and policies and perpetuated by institutions (including healthcare systems). Previous studies in Malawi suggest gender norms and masculinities (Nyirenda et al. 2006) are factors in access, attitudes and stigma (Chikovore et al. 2015; Moynihan 1998). Gender inequalities in health system could be disrupted “from within, through actions for reform and transformation, and from the outside, through progressive policies and laws and community pressure and activism” (Hay et al. 2019) involving a multi-disciplinary team of public health experts. Further studies considering both Malawian gender norms, masculinities and healthcare system inequality including geographical accessibility and means of transport are needed in order to implement effective and cost-effective interventions to increase the involvement of male partners (Manda-Taylor et al. 2017) and to achieve Sustainable Development Goals and universal health coverage (Comrie-Thomson et al. 2015).

## Conclusions

To our knowledge this is the first study evaluating the association between the socio-demographic characteristic of women in PMTCT, their knowledge, attitude and practice toward HIV/AIDS and the male partner involvement. The results showed a good level of male partner involvement compared to other studies conducted in the same context. Women’s knowledge on HIV seems to play an important role in their capacity of involving their male partners in care. Given their knowledge does not seem to result in better practice; this may suggest that other factors impact on being accompanied to the facility. Therefore, interventions targeting only women are not enough to increase the involvement of the male partners. We recommend multiple integrated component interventions with effect on health behaviour of male partners and couples, we think that addressing socio-determinants of health as socio-economic conditions, healthcare system inequality including geographical accessibility, gender norms and masculinity is of paramount importance to improve the health of all the family.

## Data Availability

All the data are available in the manuscript

## Acknowledgements

The authors wish to sincerely thank all study participants and all the healthcare staff in Malawi. Without their generous time, this study, and the results it generated, would not have been possible.

## Ethical approval

Research approval was obtained from the National Health Sciences Research Committee (Minister of Health) in Malawi (approval nr 2021).

## Abbrevations

HIV: Human Immunodeficiency Virus
AIDS: Acquired immune deficiency syndrome
DREAM: (Disease Relied through Excellent and Advanced Means)
PMTCT: Prevention Mother To Child Transmission
CHTC: Couple HIV testing and counselling
ART: Antiretroviral therapy
SSA: Sub-Saharan Africa
LTFU: lost to follow-up

